# Shared tendencies between the adoption of masking sentiment and vaccine sentiment can determine the outcome of disease spread

**DOI:** 10.1101/2024.12.02.24318343

**Authors:** Rohan S. Mehta

## Abstract

In the last two decades, the impact of human health-related behavior on the spread of the disease has increased in prominence due to the proliferation of sentiment against public health-promoting behaviors such as vaccination and mask-wearing. The recent SARS-COV-2 pandemic brought these effects into sharp focus. Recently, coupled-contagion epidemiological models have been used to study the joint impact of human behavior and disease. These models treat sentiment towards a particular public health intervention as a contagion spreading in parallel to the disease itself. In this paper, we use a coupled-contagion model to study the interaction of two different health behaviors—vaccination and mask-wearing. In particular, we study how a positive or negative association between these behaviors—e.g. a pro-vaccine person may be more likely to be pro-mask-wearing—affects disease dynamics. We find that the strength of such an association alone can determine the outcome of disease spread in the short term and the long term. In addition to studying human health-related behaviors separately, it is vitally important to understand how these behaviors are associated with each other and how these associations affect the outcomes of epidemics.

## Introduction

The effects of human behavior on disease spread are as old as modern medical interventions themselves; the first successful vaccine for smallpox was met with a of anti-vaccine sentiment from many different groups of people (1). Recent decades have seen a dramatic increase in the modeling of the effects of human behavior on disease spread (2; 3; 4; 5; 6; 7; 8). One modeling strategy that has incorporated human behavior and disease dynamics is the coupled contagion approach. In this approach, the disease and a human opinion on a health-related behavior are treated as contagions, where the spread of the opinion affects health-related behavior which then affects the spread of the disease (2; 9). Early models using the coupled contagion framework focused on either generic traits (10), disease awareness (11), or disease fear (12; 13). Mehta and Rosenberg (14) defined the term “cultural pathogen” to refer to a contagious opinion for health-related behavior that decreases the health of the carrier of the opinion. Note that this concept is distinct from the “parasitic contagion” from Hébert-Dufresne et al. (15), which describes a contagion that simultaneously harms and benefits from the presence of another contagion. The model from (14) and other recent models (e.g. 16; 17; 18) use a coupled contagion approach to study the effects of vaccine opinion on disease dynamics, and found that these opinions can determine both short- and long-term outcomes of disease spread. In the last two decades, models have also studied the effect of mask-wearing behavior on disease spread (19; 20; 21), also see citations in (8), but as far as we are aware, none aside from the bipartite network model of Qiu et al. (22) have studied this behavior under a coupled contagion framework.

In one of the first studies to use a coupled contagion model with more than one behavior and a disease, Epstein et al. (23) notably studied a model that combined a disease fear contagion with a vaccine sentiment contagion, with the result that multiple waves of infection, and changes in infection form and intensity, are signficantly affected by the characteristics of both behaviors. Recently, Teslya et al. (24) studied the effects of two mutually-exclusive, competing health opinions (one health-positive and one health-neutral) and found complex results, including the fact that both opinions can coexist and that the initial state of population opinion can affect the disease dynamics. Other models have studied the effects of multiple non-pharmaceutical interventions simultaneously (see citations in (8)) but none have studied their joint dynamics under a coupled contagion framework.

In this study, we consider a model of disease spread coupled with two pairs of competing behavioral contagions: pro-vaccine sentiment, anti-vaccine sentiment, pro-mask sentiment, and anti-mask sentiment. In addition to studying these two different health-related behaviors as cultural pathogens, we model a potential tendency of humans to adopt one type of behavior if they have already adopted another type of behavior. For example, people may be either more or less likely to adopt anti-masking sentiment if they have already adopted anti-vaccine sentiment. We study the long-term behavior of this model and establish conditions under which we expect endemic disease, endemic anti-mask sentiment, and endemic anti-vaccine sentiment. We find situations where different sentiment and disease states are potentially stable at the same time, and characterize the set of initial conditions of population sentiment in which each such state is reached. We also study the effect of the spread of masking sentiments and vaccination sentiments in the short-term on a single epidemic of a new disease. We find that the strength of the association between the two cultural pathogens can completely determine the outcome of disease spread regardless of the specific parameters of the social or biological contagions themselves, provided that the biological contagion doesn’t self-extinguish in isolation. In addition, we provide the first results of how mask-wearing sentiment acting as a cultural pathogen affects the dynamics of a disease, and show that the spread of masking behavior is overall less important than the spread of vaccination behavior unless mask efficacy is very high.

## Model

This model is a compartmental model based off of the standard Kermack-McKendrick SIR model (25). Each individual is placed into compartments from three categories: disease state, vaccine sentiment state and mask sentiment state.

The disease state is the same as in a simple SIR model with vital dynamics and vaccination: individuals can be susceptible to the disease (*S*), infected with the disease (*I*), or recovered from the disease (*R*). The R compartment also includes vaccinated individuals. Individuals enter the population as *S*, can transition from *S* to *I* by interacting with an *I* individual, and can transition from *S* to *R* by vaccination. Infected individuals can also transition from *S* to *R* by recovering from the disease.

For the vaccine sentiment state, individuals can either be pro-vaccine (*P*) or anti-vaccine (*A*). Individuals transition between the two states by interacting with individuals from the other state.

Only pro-vaccine individuals can be vaccinated (i.e. can make the *S* → *R* transition.

For the mask sentiment state, individuals can either be pro-mask (*M*), or anti-mask (*N*). Individuals transition between the two states by interacting with individuals from the other state. Pro-mask individuals exhibit a decrease in the transmission probability of the disease (i.e. the rate of the *S* → *I* transition is decreased).

Each of the 12 compartments in this model is labeled by the disease state, vaccine sentiment state, and mask sentiment state, in that order. Table 1 describes the notation for each compartment. We denote marginal variables by either one or two letters (e.g. *PM* = *SPM* + *IPM* + *RPM* and *N* = *SPN* + *SAN* + *IPN* + *IAN* + *RPN* + *RAN*). Table 2 describes the parameters used in the model.

**Table 1:** Description of model variables (compartments). The marginal variables (labeled with fewer than three letters) are sums of the 12 main variables of the model.

| Compartment | Disease State | Vaccine Sentiment State | Mask Sentiment State |
| --- | --- | --- | --- |
| SPM | Susceptible | Pro-vaccine | Pro-mask |
| SPN | Susceptible | Pro-vaccine | Anti-mask |
| SAM | Susceptible | Anti-vaccine | Pro-mask |
| SAN | Susceptible | Anti-vaccine | Anti-mask |
| IPM | Infected | Pro-vaccine | Pro-mask |
| IPN | Infected | Pro-vaccine | Anti-mask |
| IAM | Infected | Anti-vaccine | Pro-mask |
| IAN | Infected | Anti-vaccine | Anti-mask |
| RPM | Recovered | Pro-vaccine | Pro-mask |
| RPN | Recovered | Pro-vaccine | Anti-mask |
| RAM | Recovered | Anti-vaccine | Pro-mask |
| RAN | Recovered | Anti-vaccine | Anti-mask |
| SM | Susceptible |  | Pro-mask |
| SN | Susceptible |  | Anti-mask |
| PM |  | Pro-vaccine | Pro-mask |
| PN |  | Pro-vaccine | Anti-mask |
| AM |  | Anti-vaccine | Pro-mask |
| AN |  | Anti-vaccine | Anti-mask |
| S | Susceptible |  |  |
| I | Infected |  |  |
| R | Recovered |  |  |
| P |  | Pro-vaccine |  |
| A |  | Anti-vaccine |  |
| M |  |  | Pro-mask |
| N |  |  | Anti-mask |

**Table 2:** Description of model parameters.

| Parameter | Description | Range |
| --- | --- | --- |
| $b$ | Birth rate | $[0, \infty)$ |
| $r$ | Disease transmission rate | $[0, \infty)$ |
| $z$ | Reduction in disease transmission due to masking | $[0, 1]$ |
| $g$ | Recovery rate | $[0, \infty)$ |
| $v$ | Vaccination rate | $[0, \infty)$ |
| $p$ | Pro-vaccine transmission rate | $[0, \infty)$ |
| $a$ | Anti-vaccine transmission rate | $[0, \infty)$ |
| $m$ | Pro-mask transmission rate | $[0, \infty)$ |
| $n$ | Anti-mask transmission rate | $[0, \infty)$ |
| $x$ | Sentiment association | $[-1, 1]$ |

We can think of this model as consisting of three “subsystems” that interact: the disease subsystem, the vaccine sentiment subsystem, and the mask sentiment subsystem. At any given point, any individual inhabits one compartment from each subsystem. In addition, the vaccine and mask sentiment subsystems can be combined into a larger subsystem—the sentiment subsystem— that does not depend on the disease subsystem.

Figure 1A depicts the interactions between the disease, vaccine sentiment, and mask sentiment subsystems. Each of the four sentiment states acts as a simple contagion; for example, an individual transitions from proto anti-vaccine at a rate *a* upon interaction with an anti-vaccine person. Only individuals who are in the pro-vaccine state (*P*) can transition from the susceptible state (*S*) directly to the recovered state (*R*) by vaccination. Mask-wearing decreases transmission rate of the disease by a fraction *z*; we choose this simple effect of mask-wearing (following (20)) instead of a more realistic model of the efficacy of mask-wearing for the sake of simplicity.

**Figure 1:**
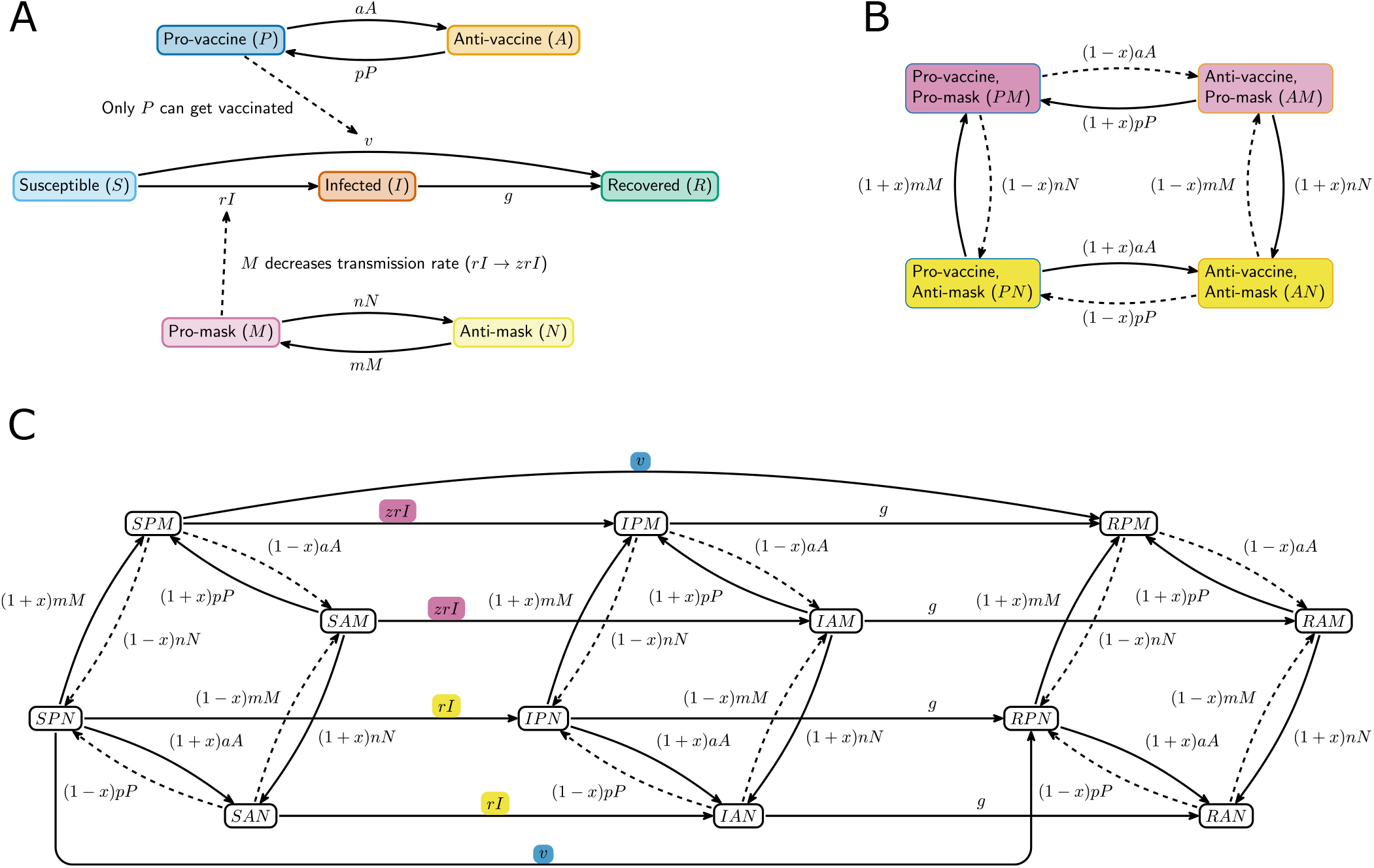
A) Interaction between sentiment subsystems and disease subsystem. Pro- and anti-vaccine and pro- and anti-mask sentiments act as simple contagions. Only pro-vaccine individuals can be vaccinated. Pro-mask individuals have a modified, decreased, probability of disease transmission. B) Interaction between sentiment subsystems. The contagion behavior of the two sentiments is mediated by the association parameter *x*. For large positive *x*, transitions labeled by solid arrows are more frequent, and for large negative *x*, transitions labeled by dashed lines are more frequent. C) Full model with all transitions between states. The two possible vaccination transitions are colored blue. The infection rates are colored by the mask sentiment; green for pro-mask (and so the infection rate is decreased), yellow for anti-mask (and so the infection rate is increased).

Figure 1B depicts how the vaccine and mask sentiment systems interact with each other. For a particular sentiment, the pro- and anti-state contagions compete with each other; the one with a higher transmission rate wins. This interaction is affected by the association parameter *x*, which ranges from −1 to 1 and describes how an individual’s tendency to exhibit a position on vaccines is similar to that individual’s tendency to exhibit a position on masks. For example, *x >* 0 means that if an individual is pro-vaccine, that individual is more likely to turn pro-mask when interacting with a pro-mask individual than if the individual were anti-vaccine. An individual is also more likely to turn pro-vaccine if the individual is pro-mask. The opposite occurs for *x <* 0; an individual is more likely to turn pro-vaccine if that individual is anti-mask and more likely to turn anti-vaccine if pro-mask. When *x* = 0, there is no association between behaviors, so the two contagions are independent.

Figure 1C depicts all the possible transitions between all the compartments of the model. The four states that correspond to a particular disease state and all possible sentiment state combinations can each transition among each other via the four different sentiment contagions. The four susceptible states can become infected, but the infection rates for *SPM* and *SAM* (the pro-mask states) are decreased compared to those for *SPN* and *SAN* (the anti-mask states) by a factor of *z*. We use the standard assumption that only one state transition can happen at once (so that e.g. the transition *SPM* → *IAN* is not allowed.)

Eq. 1 shows the full system of equations corresponding to the model from Figure 1C. Note that all variables must sum to 1, so we can omit one variable (*RAN*) from most further analysis.

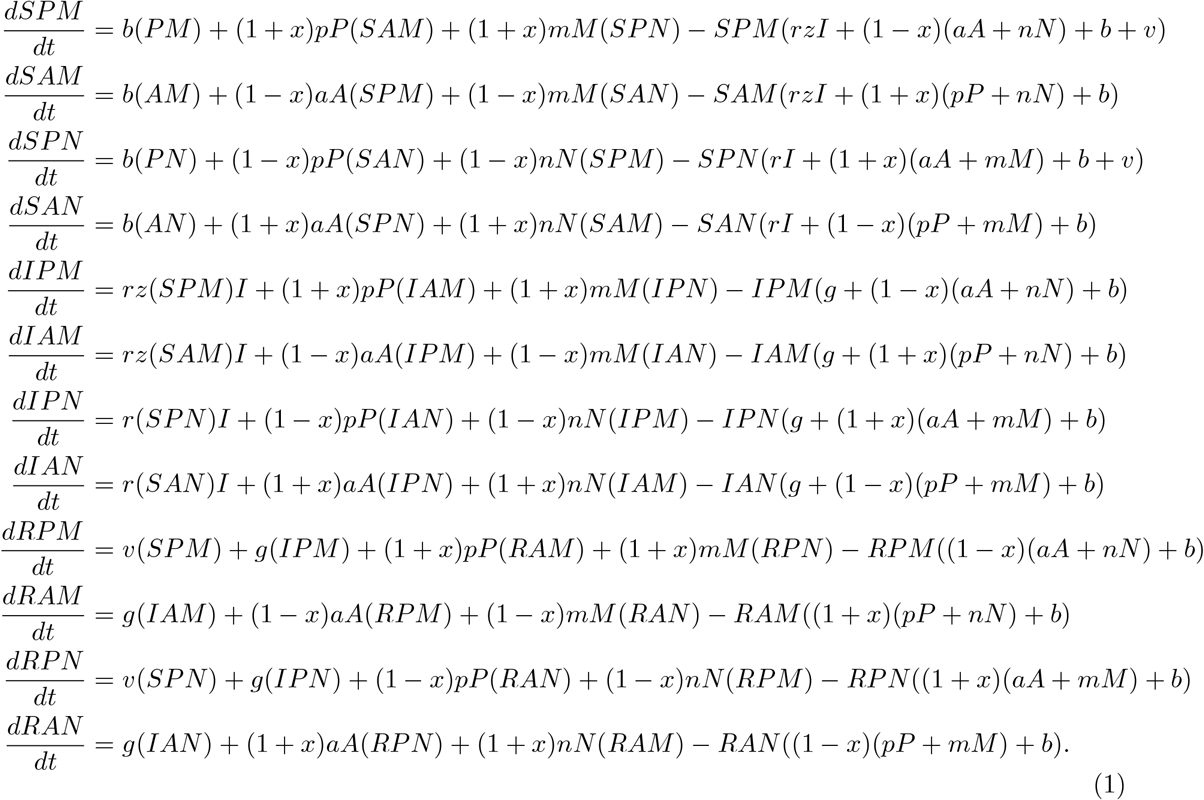

## Results

### Long-Term dynamics

#### Sentiment Equilibria

In the long run, this system reaches one of several possible stable equilibria. The dynamics of the sentiment subsystem are independent of the dynamics of the disease subsystem (but not vice versa). The sentiment subsystem is

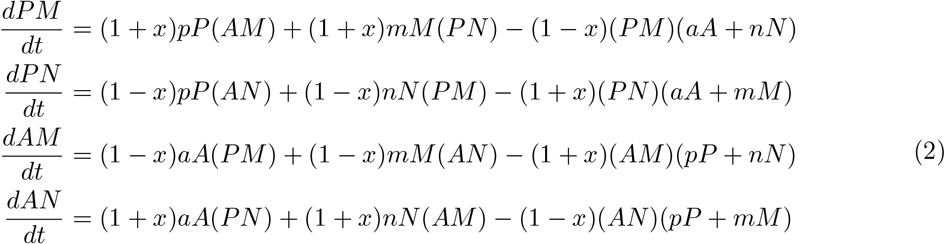

There are four sentiment subsystem equilibria that are nontrivial and do not rely on specific equalities in parameter values, each corresponding to one particular sentiment type (*PM, PN, AM*, or *AN*), taking over the population. See Appendix for details.

To express the stability conditions for these equilibria, we use the following two quantities:

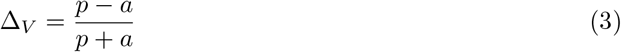

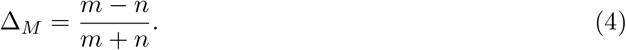

Δ_*V*_ (eq. 3) is the relative transmissability or persuasiveness of pro-vaccine sentiment compared to anti-vaccine sentiment, and Δ_*M*_ (eq. 4) is the equivalent value for masking sentiment. Table 3 summarizes the stability conditions for the four possible equilibria. From Table 3 and the knowledge that −1 ≤ *x*, Δ_*V*_, Δ_*M*_ ≤ 1, we can deduce that ranging *x* from -1 to 1 will yield all possible sentiment states as stable at some point, at least in a situation where bistability is possible (i.e. the bottom two columns of Table 3).

**Table 3:**
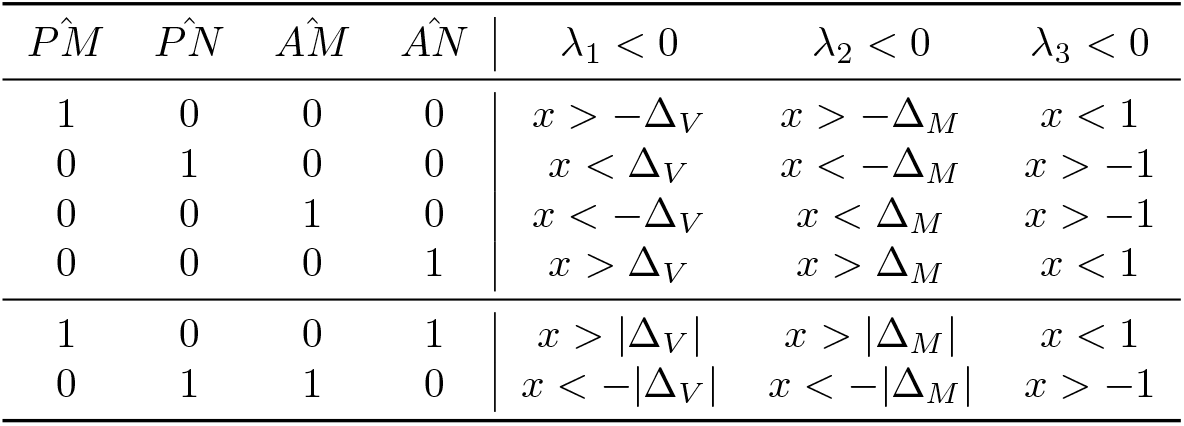
Equilibria and stability conditions for the sentiment subsystem. Note that the last two rows are for bistability; the population is not ever simultaneously in a state where 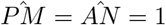 or 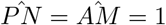.

The effect of *x* can be summarized by stating that a strong enough association can override existing relationships between sentiments to “drag” a weaker position into dominance if the corresponding position in the other sentiment is strong enough. For example, *PM* can be stable even if anti-vaccine or anti-mask sentiment is dominant (Δ_*V*_, Δ_*M*_ *<* 0) so long as *x* is large enough. If one one of those “anti” states is dominant, “large enough” only depends on the strength of that state. The conditions in Table 3 leave the possibility open for multiple equilibria to be stable at once. The first possible situation involves a very large positive *x*, in which case the association can overpower sentiment transmission and “drag” the double-pro (*PM*) or double-anti (*AN*) state into dominance. The second possible situation involves a very large negative *x*, which has the opposite effect and can drag one of the “mixed” states (*PN* or *AM*) into dominance.

Because there is complete symmetry between the two sentiments in the sentiment subsystem, we can choose exactly one of each option for several choices without loss of generality as an illustrative example of the potential for bistabilty: we focus on the case where the double-pro sentiments *PM* and *AN* are possible, we let Δ_*V*_ *>* Δ_*M*_ *>* 0, and range over a variety of initial conditions to see when which cases *AN* dominates vs *PM*. In this case, we require *x* to be large but less than 1, and we expect that we are in the basin of attraction of the *AN* equilibrium more when we start close to the equilibrium (i.e. *AN*_0_ ≈ 1) and when *x* is larger. Because Δ_*V*_, Δ_*M*_ *>* 0, the two “positive” sentiment contagions are stronger than the “negative” ones, so the pair *PM* should dominate under more initial conditions.

This reasoning is demonstrated in Figure 2, where we fixed *p* = 0.8, *a* = 0.2, *m* = 0.6, and *n* = 0.4. Thus, Δ_*V*_ = 0.6 and Δ_*M*_ = 0.2. We ranged *x* from 0.6 to 0.95. These parameters satisfy the condition in Table 3 for bistability of *PM* and *AN*. For simplicity, we fixed *PN*_0_ = *AM*_0_ = 0.01 and ranged *PM*_0_ from 0 to 1 in intervals of 0.01. In this case, *AN* = 1 when *AN*_0_ is large enough (and *PM*_0_ is small enough). Figure 2 shows the minimum value of *AN*_0_ such that 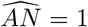 for a range of values of *x* such that bistability is possible. We see that as 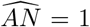 increases, it becomes easier for the unfavored pair *AN* to dominate over the favored pair *PM*, demonstrating that association alone between behaviors is sufficient to switch the outcome of the dynamics.

**Figure 2:**
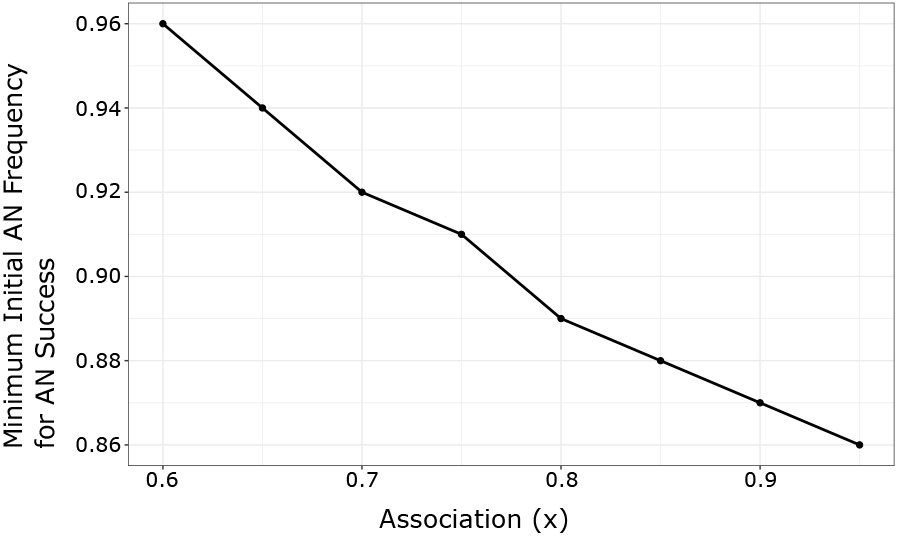
The minimum required initial frequency for *AN* to dominate over *PM* when both are potentially stable decreases with increasing association *x*. Thus, a stronger association between behaviors can change a situation from *PM* to *AN*, potentially dramatically shifting the outcome of the disease spread. Parameters used in these simulations are *p* = 0.8, *a* = 0.2, *m* = 0.6, *n* = 0.4, *x* ∈ [0.6 − 0.95], and *PM*_0_ ∈ [0, 1].

#### Disease Equilibria

We can use the sentiment subsystem results to find the different disease subsystem equilibria. The disease subsystem is

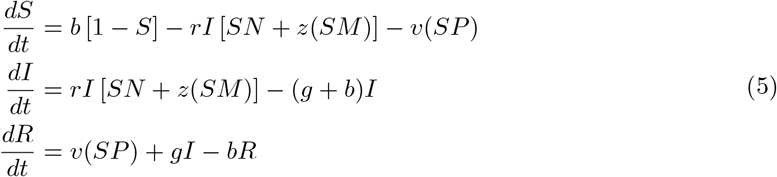

First, we define the standard epidemiological quantity

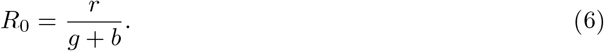

There are two possible options for the equilibrium value of infected individuals *Î*: either *Î* = 0 and the disease is extinct in the long term or *Î >* 0 and the disease is endemic. Each of these options can combine with all four of the sentiment subsystem equilibria to create 8 possible situations. Table 4 lists these situations with the corresponding equilibrium values of the variables. As seen in Table 4, the presence of both vaccination and masking decreases endemic disease.

**Table 4:** Disease equilibrium values for each sentiment equilibrium scenario. Note that for each scenario, only one pair of sentiment states exists, so there are only three potential nonzero states. The recovered state can be obtained by subtracting the susceptible and infected states from 1.

| Sentiment Equilibrium | $\hat{S}$ | $\hat{I}$ | Stability Condition |
| --- | --- | --- | --- |
| $\widehat{PM} = 1$ | $\frac{b}{v+b}$ | 0 | $R_0 < \frac{1}{z} \left(1 + \frac{v}{b}\right)$ |
| $\widehat{PM} = 1$ | $\frac{1}{zR_0}$ | $\frac{b}{g+b} \left[1 - \left(1 + \frac{v}{b}\right) \frac{1}{zR_0}\right]$ | $R_0 > \frac{1}{z} \left(1 + \frac{v}{b}\right)$ |
| $\widehat{PN} = 1$ | $\frac{b}{v+b}$ | 0 | $R_0 < 1 + \frac{v}{b}$ |
| $\widehat{PN} = 1$ | $\frac{1}{R_0}$ | $\frac{b}{g+b} \left[1 - \left(1 + \frac{v}{b}\right) \frac{1}{R_0}\right]$ | $R_0 > 1 + \frac{v}{b}$ |
| $\widehat{AM} = 1$ | 1 | 0 | $R_0 < \frac{1}{z}$ |
| $\widehat{AM} = 1$ | $\frac{1}{zR_0}$ | $\frac{b}{g+b} \left[1 - \frac{1}{zR_0}\right]$ | $R_0 > \frac{1}{z}$ |
| $\widehat{AN} = 1$ | 1 | 0 | $R_0 < 1$ |
| $\widehat{AN} = 1$ | $\frac{1}{R_0}$ | $\frac{b}{g+b} \left[1 - \frac{1}{R_0}\right]$ | $R_0 > 1$ |

Figure 3 displays how the critical *R*_0_ (aka the value at which *R*_0_ must exceed for the disease to be endemic) varies with the association parameter *x* and the sentiment subsystem equilibrium in the case where Δ_*V*_ *<* Δ_*M*_ *<* 0 (so each anti-contagion is stronger than each pro-contagion, and anti-vaccine sentiment is stronger than anti-mask sentiment). For illustrative purposes, we chose *b* = 0.02, *v* = 0.14, and *z* = 0.3. The critical *R*_0_ is lowest when both anti-sentiments dominate 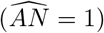 and highest when both prosentiments dominate 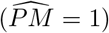. The multiplicative nature of the joint effect of masking and vaccination is visible in how much higher the critical *R*_0_ is for 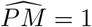 than for both 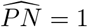 and 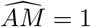, where only one prosentiment is dominant.

**Figure 3:**
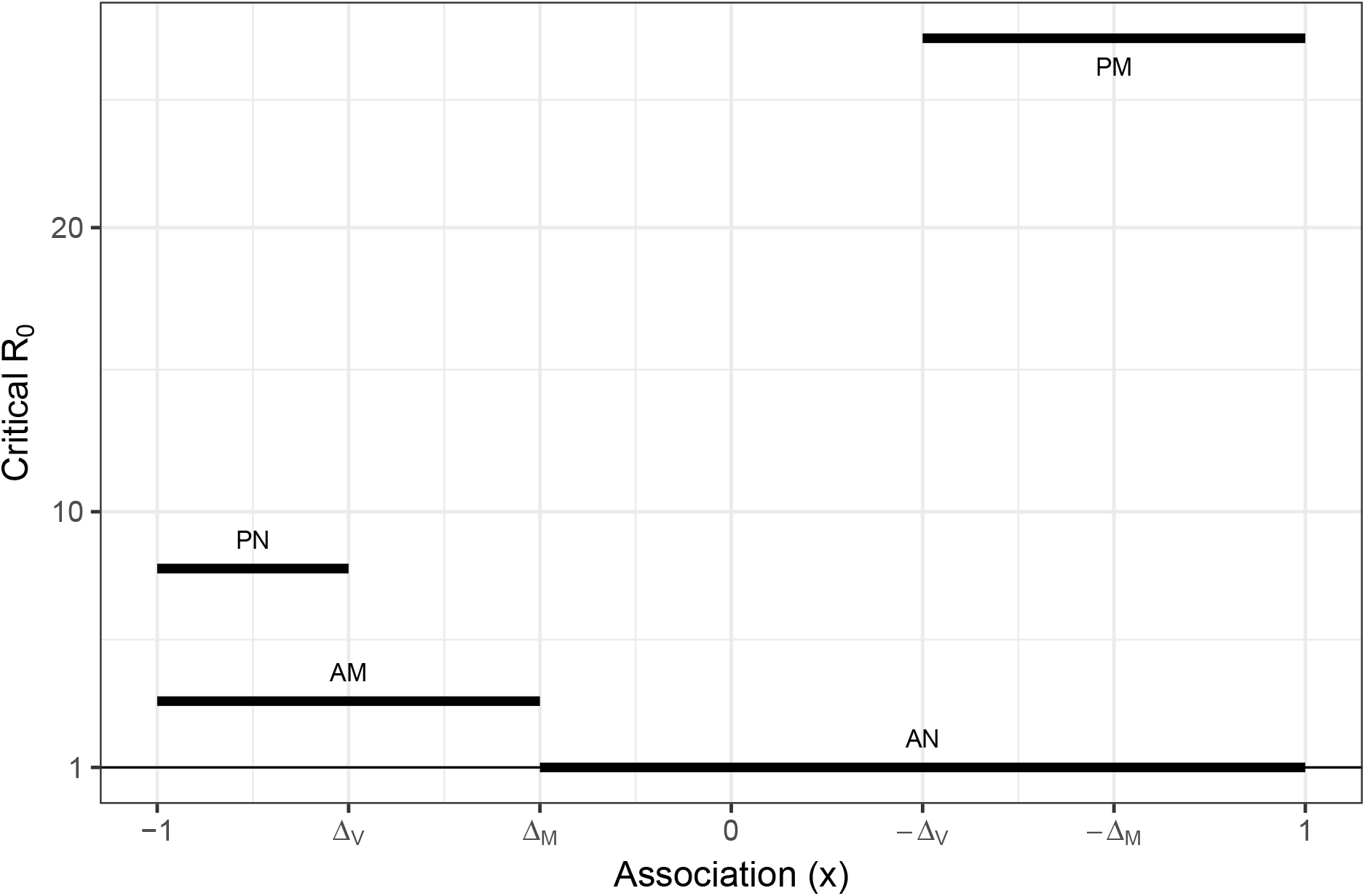
The critical value of *R*_0_ that a disease must exceed to be endemic is highest when both pro-sentiments are dominant and lowest when both anti-sentiments are dominant. The association parameter *x* determines which sentiment state is dominant, and any dominant sentiment state can be obtained with a particular value of *x*. Critical *R*_0_ values are obtained from Table 4. The horizontal lines indicate critical values of *R*_0_, labeled by which sentiment state is dominant. Δ_*V*_ and Δ_*M*_ are defined in eq. 3 and eq. 4, respectively.

### Example

Figure 4 displays the equilibrium value of disease (*Î*) for each sentiment equilibrium, a range of values of *z* and *v* and disease parameters *b* = 0.02, *r* = 2.14, *g* = 0.014, *R*_0_ ≈ 63 (chosen for demonstrative purposes). In a situation with a highly contagious disease, in order to eliminate the disease in the long-term, it is critical for pro-vaccine sentiment to dominate. Mask-wearing sentiment has a comparatively smaller effect on endemic disease. The effec of the two sentiments combined is multiplicative.

**Figure 4:**
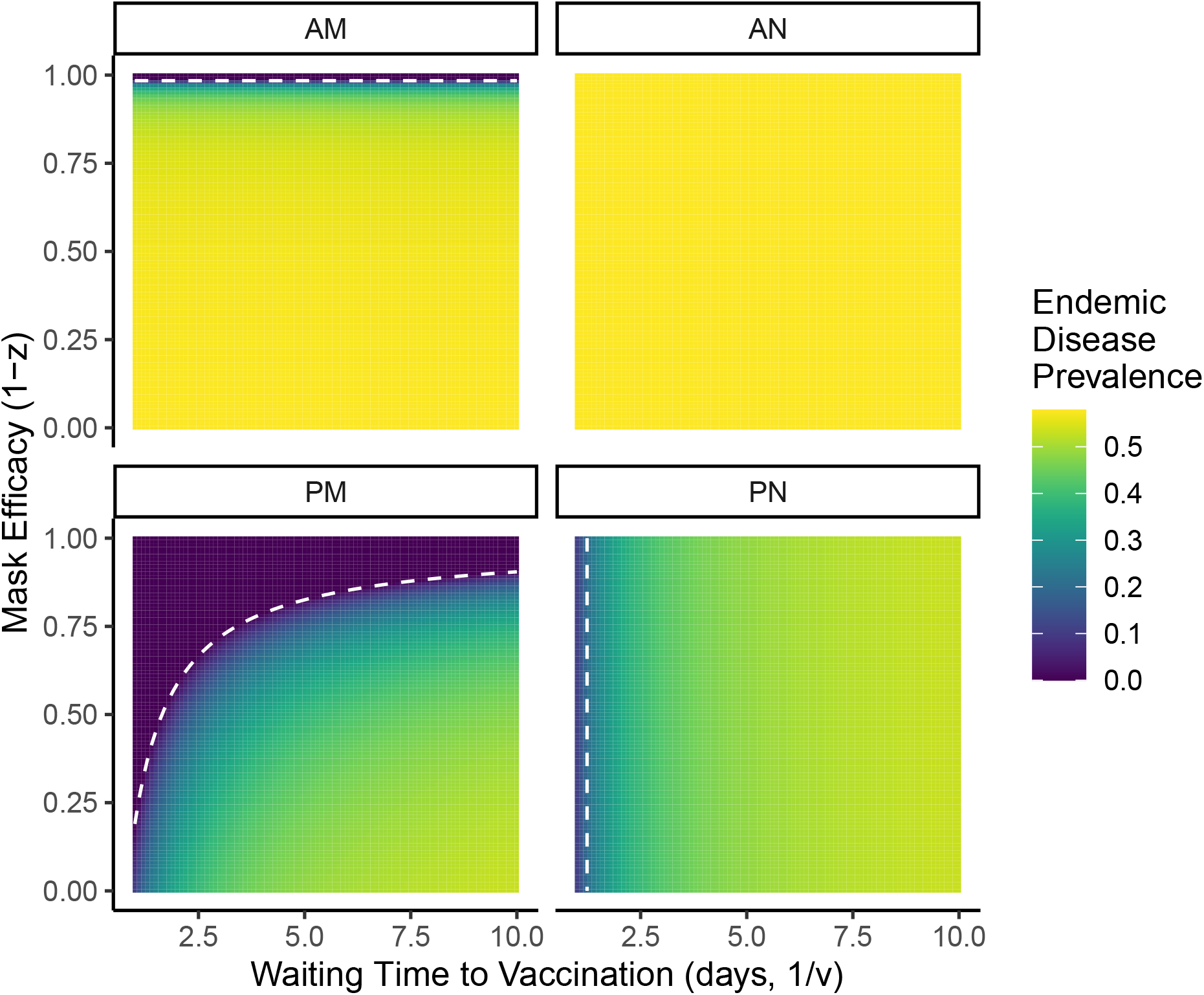
Endemic disease prevalence is affected by which sentiment state is dominant. For the two anti-vaccine sentiment states, mask efficacy makes little difference on endemic disease. For the two pro-vaccine states, vaccination rate and mask efficacy have strong effects, leading to situations where sentiment presence can affect disease endemism. The white dashed lines represent the boundary between disease endemism at equilibrium and disease extinction at equilibrium

### Short-Term Dynamics

Here we study the effect of associated masking and vaccination behavior on a single epidemic of a new disease. We assume that a population is generally pro-vaccine and pro-mask with respect to this disease. The only non-susceptible, non-pro-vaccine, and non-pro-mask individuals in the population are one infected individual, one anti-mask individual, and one anti-vaccine individual, with every other state of each of these individuals pro-sentiment and/or disease susceptible. Our initial condition is

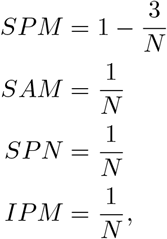

and everything else zero. For our parameters, we use disease parameters representative of measles (as used in (14)), with *b* = 0 so that we look at a single instance of potential disease spread over a short time scale. For our sentiment parameters, to simplify things, we set *p* + *a* = *m* + *n* = 1 and vary *x* ∈ [−1, 1], *z* ∈ [0, 1], and *v, p, m* ∈ [0, 1].

Figure 5 displays the short-term dynamics of the disease in this system, summarized by two measurements. First, we calculate the maximum fraction of the population infected (i.e. the height of the epidemic peak). Second, we calculate the total area under the infected curve divided by its maximum possible value of 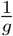 (aka the “total infection load”).

**Figure 5:**
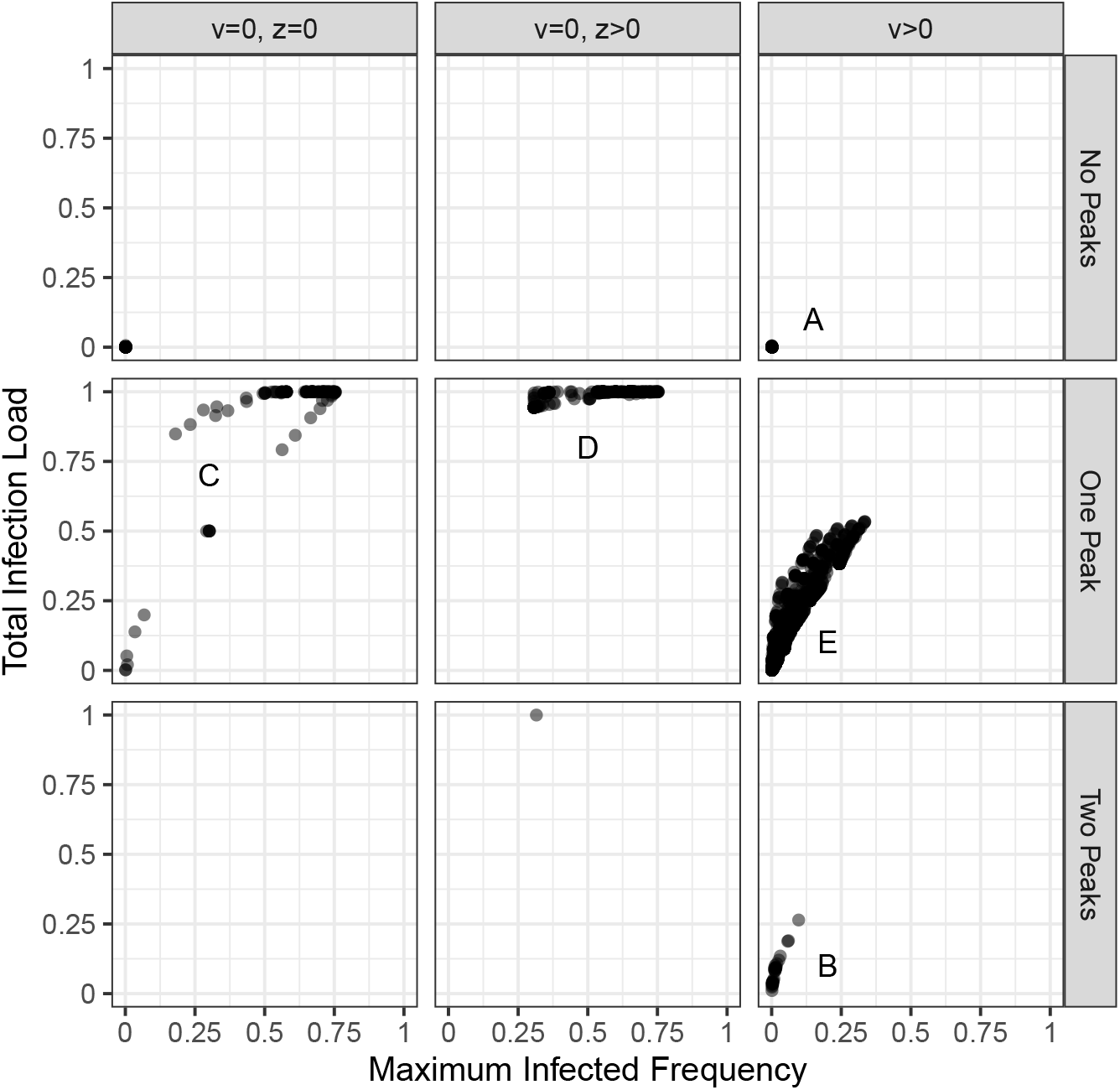
Short-term dynamics of this system, described by the maximum infected frequency at the peak of the epidemic and the total infection load (the integral of the infected frequency divided by its maximum). We divide the results by the number of epidemic peaks. We also divide the results into *v* = 0 and *v >* 0; when *v >* 0 both measured variables are much lower than when *v* = 0. We further divide the *v* = 0 case into *z* = 0 and *z >* 0; when masking is not perfectly effective and there is no vaccination, it is very difficult to avoid maximum infection load. Labels A-E refer to the panels in Figure 9, which shows example trajectories in each of the labeled panels.

Observation of the results suggested three different categories related to masking and vaccination effectiveness. The first is when *v* = 0 and *z* = 0; masks are perfectly effective and there is no vaccination. The second is when *v* = 0 and *z >* 0; masks are imperfectly effective and there is no vaccination. Finally, the third category is when and *v >* 0; there is vaccination present. We also divide the results by the number of epidemic peaks observed in the simulation.

When there is no vaccination (i.e. *v* = 0), most of the results involve a total infection load at its maximum possible value 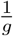, equivalent to starting the entire population as infected and then having the infected population decay exponentially at a recovery rate of −*g*) and maximum infection frequency of at least 30%. When masking is imperfectly effective, the total load remains very high, whereas decreased total load is possible when masking is perfectly effective. When there is vaccination, the total infection load stays below 0.5, and the maximum infection frequency stays below 40%. We explore these situations in more detail below.

Another aspect of the infection dynamics is how many local maxima occur. For this parameter sweep, we observe three possible situations: no peaks (aka monotonic decrease from the initial frequency of 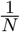, one peak (a single epidemic), and two peaks (two epidemics, usually with the second much stronger than the first, occurring due to changes in sentiment dynamics).

### No Epidemic Peaks

When there is no epidemic peak, the infected frequency starts at the initial frequency of 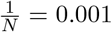 and then monotonically decreases. Thus, the maximum infection frequency is 0.001. The total infection load thus depends on how quickly the infected frequency decreases. When there is no vaccination and masking is perfectly effective (*v* = 0,*z* = 0), the primary driver of this decrease is the spread of mask-wearing in the population. More masks means slower transmission and therefore faster decay. One might think that the spread of vaccine sentiment has no effect when *v* = 0, but if the behaviors are associated, the spread of either pro- or anti-vaccine sentiment will speed up or slow down the decay rate of the infected population, with the direction of the effect depending on the direction of the association.

Figure 6 shows the effects of sentiment transmission and sentiment association on the total infection load when there are no peaks (or epidemics). When there is no vaccination (*v* = 0), the pro-vaccine sentiment spread can increase pro-mask sentiment spread when *x >* 0, leading to almost no infection load. When *x <* 0, pro-vaccine sentiment spread increases anti-mask sentiment spread, so it increases infection load. Masking always decreases infection load.

**Figure 6:**
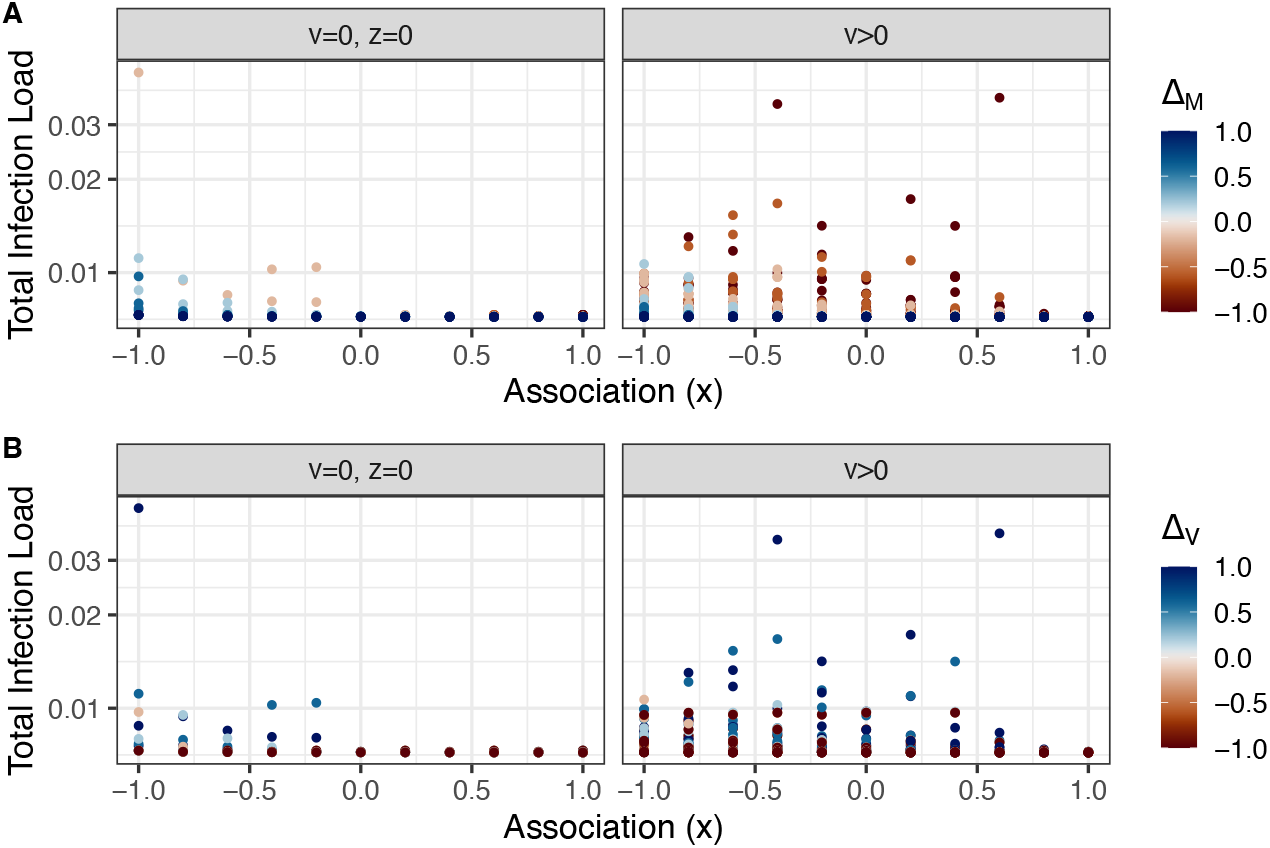
When there is no epidemic, the total infection load is affected by the association between the two sentiments for (A) mask sentiment, and (B) vaccine sentiment. In general, the spread of pro-mask sentiment decreases infection load (as infecton load is low for most blue values in A). When *x <* 0, the spread of pro-vaccine sentiment decreases the spread of pro-mask sentiment, increasing the disease load (blue values in B tend to be higher when *x <* 0).

When there is vaccination (*v >* 0), the effect of vaccine sentiment transmission is messier, while the effect of mask sentiment transmission is qualitatively similar to when *v* = 0, just more dramatic because the spread of masking sentiment influences the spread of vaccine sentiment and therefore vaccination rate as well.

A necessary (but not sufficient) condition for there to be no peaks is for 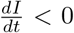 at *t* = 0 (the condition would be sufficient if it is true for all *t*). In general, if we assume that there is one anti-mask and one infected individual in a population of size *n*, this condition is equivalent to

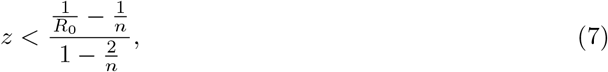

which, using our *R*_0_ = 15.29 and *n* = 1000 yields *z <* 0.065. In general, for a new disease that is approximately as infectious as measles where sentiment is almost entirely pro-mask, mask efficacy has to be almost perfect to prevent an epidemic. If there is existing immunity in the population, the condition becomes much less stringent; for a particular masking efficacy *z*, the maximum initial susceptible population that still leads to no epidemic is

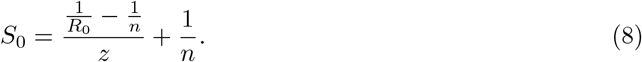

### Two Epidemic Peaks

The phenomenon of two epidemic peaks occurs almost always only when *v >* 0 (Figure 5). In addition, there is a special condition that all the two-peak simulations share:

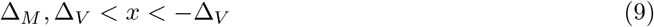

This condition requires Δ_*V*_ *<* 0, so anti-vaccine sentiment beats pro-vaccine sentiment. Using the conditions in Table 3, this condition implies *AN* as the dominant sentiment state in the long term. Thus, even if pro-mask sentiment is stronger than anti-mask sentiment (Δ_*M*_ *>* 0), a large enough association and a strong enough anti-vaccine sentiment can “drag” the anti-mask sentiment into success.

As this condition (eq. 9) summarizes the major effects of the sentiment parameters on the existence of multiple epidemics, in Figure 7 we plot the distribution of *v* and *z* values that simulations which exhibit two peaks have. We also plot the *v* and *z* distributions for simulations under the special condition (eq. 9) but with one peak, and finally the *v* and *z* distributions for every other simulation. Given the special condition (eq. 9), two peaks occur with low vaccination rate and high mask efficacy as compared to other simulations whose parameters satisfy the special condition.

**Figure 7:**
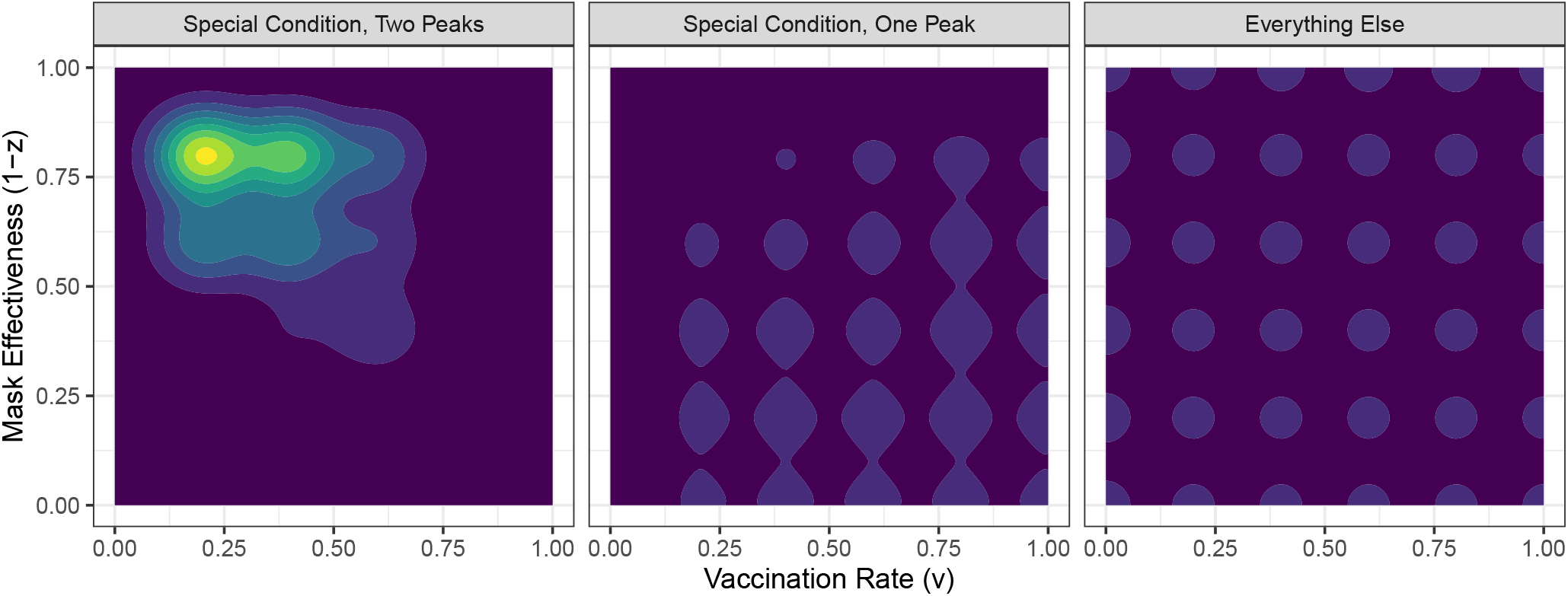
Two epidemic peaks occur for a specific condition describing sentiment dynamics that lead to pro-vaccine sentiment dominating (eq. 9) as well as high mask efficacy and low vaccination rates. The left panel shows the joint vaccination rate and mask efficacy distribution for parameter sets that lead to two epidemic peaks. The middle panel shows this distribution for all other parameter sets that satisfy eq. 9, the “Special Condition”. Finally, the right panel shows this distribution for all other parameter sets. The color scheme increases by 1 at each gradient, so the darkest color is 0 parameter sets and the lightest color is 8 parameter sets. The gridlike pattern seen in the latter two panels is due to the fact that the parameter sets are chosen in a grid search.

### One Epidemic Peak

Finally, for the large, relatively cohesive set of simulations where *v >* 0 and the number of peaks is 1, we can summarise the effects of various parameters as their effects on maximum infection frequency; the relationship between that value and total infection load is relatively strong in this case due to the fact that vaccination rate strongly controls the value.

Figure 8 displays the relationship between the maximum infection load and the model parameters. The strongest effects are of vaccination rate and mask efficacy (Figure 8A); lower vaccination rates and lower mask efficacies increase the peak height. There is also a small interactive effect of association and the relative spread of vaccine sentiment (Figure 8B); for more negative associations, the spread of anti-vaccine sentiment can lead to a higher peak.

**Figure 8:**
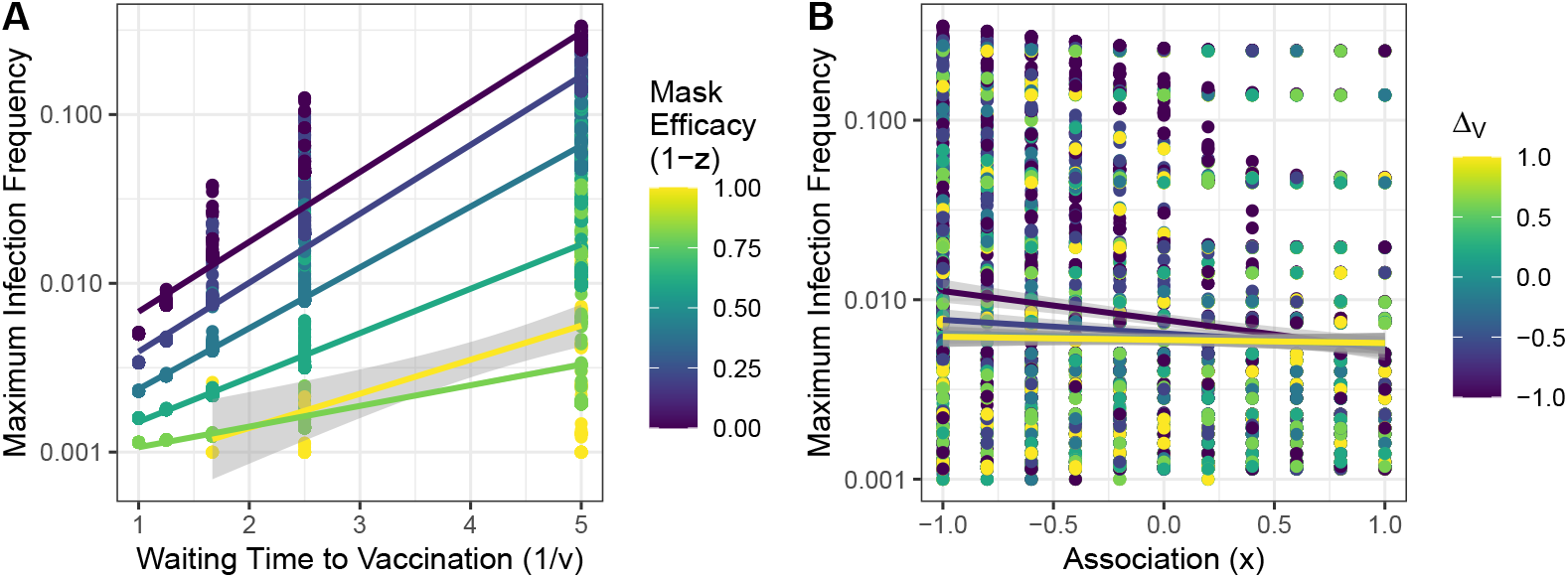
Model parameters can affect the height of the epidemic peak. A) Decreasing vaccination rate and/or mask efficacy increases peak height. B) Increasing the spread of anti-vaccine sentiment increases peak height, but only when sentiment association is more negative. This effect is due to the fact that our initial condition has almost all individuals pro-vaccine and pro-mask; so a strong negative sentiment association is needed for effective anti-vaccine or anti-mask sentiment spread.

Figure 9 contains example trajectories for selected cases in Figure 5. Parameters used in these simulations are found in Table 5.

**Table 5:** Parameters used for the simulations in Figure 9. Parameters are grouped in terms of their role in the model. Δ_*V*_ is from (eq. 3) and Δ_*M*_ is from (eq. 4).

| Panel | $v$ | $z$ | $p$ | $a$ | $m$ | $n$ | $x$ | $\Delta_V$ | $\Delta_M$ |
| --- | --- | --- | --- | --- | --- | --- | --- | --- | --- |
| A | 0.2 | 0 | 0.2 | 0.8 | 0.4 | 0.6 | 0.2 | -0.6 | -0.2 |
| B | 0.2 | 0.2 | 0.2 | 0.8 | 0.4 | 0.6 | 0 | -0.6 | -0.2 |
| C | 0 | 0 | 0.6 | 0.4 | 0.6 | 0.4 | -1,-0.4 | 0.2 | 0.2 |
| D | 0 | 0.2 | 0.8 | 0.2 | 0.4 | 0.6 | $\pm 0.6$ | 0.6 | -0.2 |
| E | 0.2 | 0.6 | 0.2 | 0.8 | 0.4 | 0.6 | $\pm 0.8$ | -0.6 | -0.2 |

**Figure 9:**
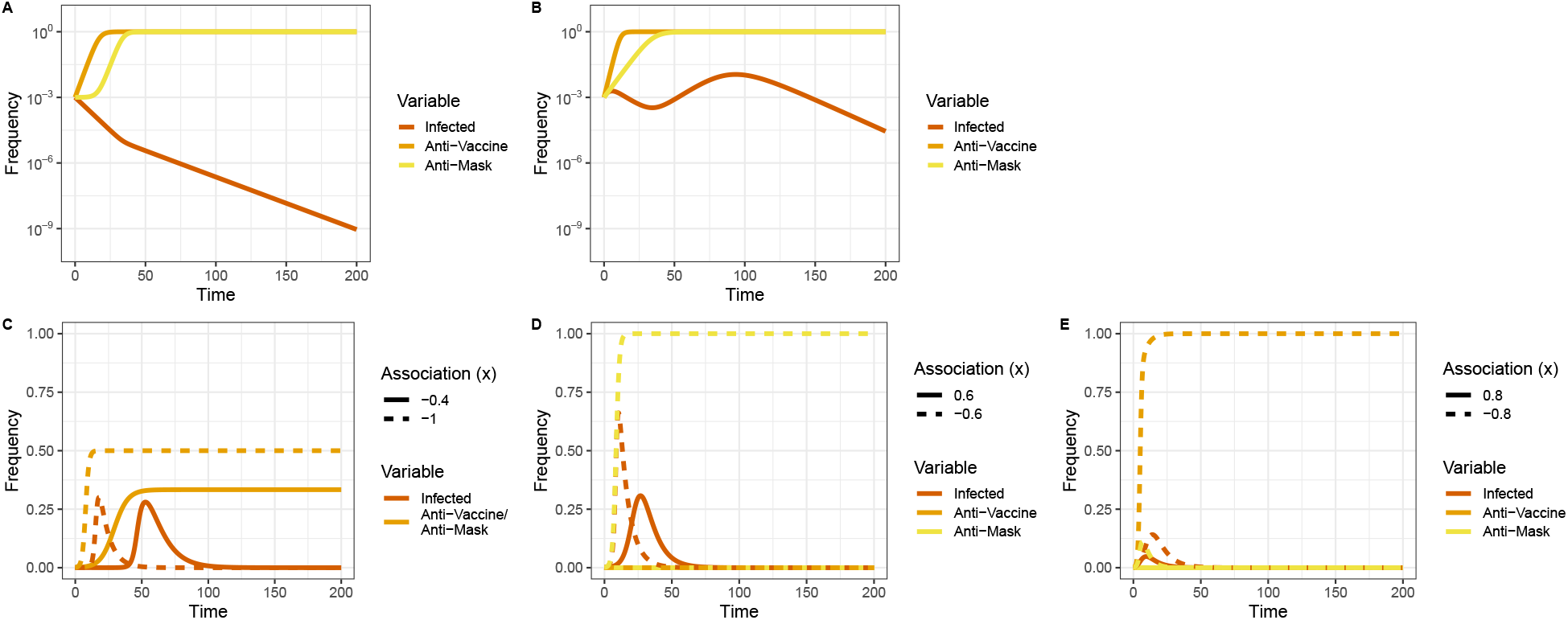
Example short-term dynamics of the model with specific panels chosen from their labeled locations in Figure 5. A) An example trajectory with no disease peak. Both anti-mask and anti-vaccine sentiment dominate, but vaccination and masking are initially too effective for the disease to gain a foothold. B) An example trajectory with two disease peaks. In this case, vaccination and masking are weak enough that the rapid spread of anti-vaccine sentiment (followed by the spread of anti-mask sentiment) allows for the disease to return after an initial epidemic. C-E) Example trajectories where there is one epidemic peak from each column in Figure 5. In these examples, the association parameter *x* is varied between a more negative number and a more positive number. In each case, the more negative number leads to a stronger epidemic, due to the fact that initially almost everyone in the population is pro-vaccine and pro-mask, and therefore negative association is required for one of the anti-mask/vaccine sentiments to spread effectively. The cases in C and D have a vaccination rate of 0, and so the spread of masking sentiment is key. In C, the spread of both are identical so they overlap. In E, however, the vaccination rate is positive, resulting in both lower overall epidemic peaks and also the spread of anti-vaccine sentiment driving the stronger epidemic. Parameters used for these simulations are found in Table 5.

The first example trajectory (Figure 9A) depicts a situation where there is no epidemic, but the anti-vaccine and anti-mask sentiments dominate the population. This example was chosen to demonstrate that for a new disease, the conditions for no epidemic to occur can be so extreme that the spread of cultural pathogens has no effect. In this case, the extreme condition is that *z* = 0; that is, masks are perfectly effective. Note that vaccination exists in this example (*v* = 0.2, see Table 5).

The example trajectory for two peaks (Figure 9B) demonstrates that the second epidemic peak can in fact be stronger than the first, due to the successful transmission of anti-vaccine and anti-mask sentiments as well as low vaccination rates. In this example, there is no association between the sentiments (*x* = 0). This value satisfies the condition in eq. 9, reflecting the fact that the absolute association value is not as important as the relationship between the association between the sentiments and the strength of the sentiment transmission itself.

The final three examples (Figure 9C-E) are for disease trajectories with one peak (epidemic). The next example (Figure 9C) shows two situations where *v* = *z* = 0 with one epidemic peak that have very similar maximum infected frequencies, very different total infection loads, and differ only in the value of *x* (*x* = −1 or *x* = −0.4).

Because there is no vaccination and masking is perfectly effective, the spread of anti-mask sentiment effectively serves as introducing new susceptible individuals into the population. When anti-mask sentiment spreads quickly, the infection spreads quickly and “burns out” fast. When anti-mask sentiment spreads slowly, the infection starts slow but is continuously replenished with new susceptible individuals so it drags on for a longer period of time.

In this particular case, Δ_*M*_ = Δ_*V*_, so both sentiments should have the same trajectory as they start with the same frequency 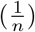. The magnitude of *x* affects the rate of spread; for *x* = −1, the sentiments spread faster, and for *x* = −0.4, the sentiments spread slower.

Note that “Total Infection Load” does not equal the total number of individuals infected. In fact, for the *x* = −1 case, about 93% of individuals in the population are infected at some point, but for the *x* = −0.4 case, only about half of the population are.

The example for *v* = 0 and *z >* 0 (Figure 9D) demonstrates the ability for sentiment association to lead to very different maximum infection frequency while maintaining the same total infection load (close to as high as possible). Because there is no vaccination, the spread of masking sentiment is key. Because almost everyone in the population is initially pro-vaccine or pro-mask, the initial sentiment spread is driven by the pro- sentiments. Thus, anti-mask sentiment can only initially spread if it is dragged along by the spread of *pro-vaccine* sentiment, which occurs when the association is negative (*x* = −0.6) but not when the association is positive (*x* = 0.6).

Finally, the example for *v >* 0 (Figure 9E) demonstrates that when there is vaccination, increasing anti-vaccine sentiment can make the disease spread faster. Also, the overall presence of the disease is generally lower with vaccination, as one would expect. This example also demonstrates the continued pattern that, due to the initial condition setup, decreasing the association parameter *x* increases the ability of anti-vaccine sentiment to transmit successfully.

## Discussion

We have developed a model of two cultural pathogens: one for mask-wearing sentiment, and one for vaccine sentiment, that affect the transmission of a disease. Our focus was to see how a tendency to associate one cultural pathogen with another affects the disease dynamics. We modeled this tendency using a parameter that affected the rate of transmission of a social contagion based on the current sentiment state of the receiver. A positive value of this parameter indicated that there was an association between “like” sentiments (e.g. pro-vaccine people are more receptive to pro-mask sentiment, and anti-vaccine people are more receptive to anti-mask sentiment), and a negative value of this parameter indicated that there was a an association between “unlike” sentiments (e.g. pro-vaccine people are more receptive to anti-mask sentiment, and anti-vaccine people are more receptive to pro-mask sentiment). Negative values of this parameter may seen unlikely to exist in reality, but it is quite simple to come up with scenarios in which a negative value might exist, at least locally. For instance, people may be anti-vaccine because they are worried about the vaccine, but are still concerned about the disease itself, and therefore would be more likely to adopt other behaviors that would prevent them from getting the disease, such as masking. Likewise, people who are vaccinated may be less likely to want to wear a mask in addition, as masks are uncomfortable and the protection from the vaccine may be judged to be sufficient.

We found that such a tendency can have a dramatic effect on the outcome of disease spread, to the extent that it can entirely determine the outcome of the spread of a disease. In the long term, for any particular social contagions, there exist critical values of the association such that any outcome of social contagion transmission can be reached. Some of these values can be quite extreme and require a very strong association between the two traits, but in principle they exist. Thus, not only can the dynamics of the social contagions themselves determine the outcome of disease spread, but the association between the social contagions can as well.

We also found a wide variety of short-term effects of the joint spread of associated vaccine and mask sentiments. The spread of these sentiments can affect how quickly a disease dies out without an epidemic, the overall strength of a single-peaked epidemic, and the possibility of multiple epidemic peaks. In addition, the association between the two sentiments can have complex effects on the spread of the sentiments and the spread of disease. If the association is strong enough, sentiments that normally would not spread in isolation can still spread in the presence of a more favorably-spreading associated sentiment, leading to changes in the existence, strength, and timing of epidemics.

The association parameter can be considered in the context of a “parasitic contagion” defined by (15), in which a contagion can increase its own spread by using another contagion. Depending on the sign of the association, different pairs of contagions in this model can be considered as having a parasitic relationship. This framework is useful for classifying social contagions that may behave in similar ways despite affecting different behaviors.

One shortcoming of this model is that it is unwieldy. There are 12 compartments, and computing the eigenvalues of the full Jacobian matrix has little utility. Future research that models multiple behaviors at once in conjunction with a disease may be better served by agent-based models where less bookkeeping is required. Another shortcoming of this model is the simplistic treatment of the effect of mask-wearing on disease transmission. It is well-known that mask-wearing has asymmetric effects on disease transmission (20). Our simplified choice was made purely for modeling convenience and simplicity; there is certainly room for future work that treats mask-wearing sentiment as a social contagion and uses more realistic more realistic modeling of disease transmission with masking.

## Data Availability

This study contains no data.

## Acknowledgements

We would like to acknowledge Noah Rosenberg for initializing interest in this topic, and Rahul Dholakia for providing workspace resources.

## A Appendix

To simplify the analysis of this system, we can study the equilibria and stability conditions of the disease and sentiment subsystems obtained by tracking marginal variables.

### A.1 Disease Subsystem Equilibria

If we track the disease states *S* and *I* (noting that *R* = 1 − *S* − *I*), we obtain

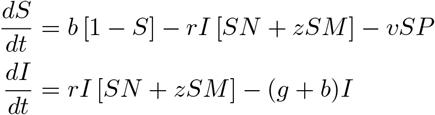

Let the dummy variables 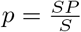 and 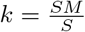

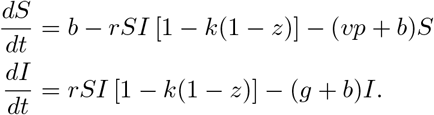

As with the standard SIR model, there are two choices for *Î*. First, *Î* = 0:

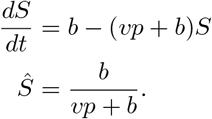

Second, *Î >* 0. First, we solve for *Ŝ*:

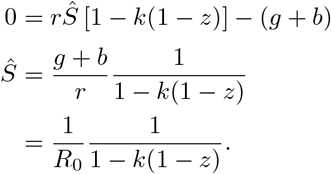

Then, we solve for *Î*:

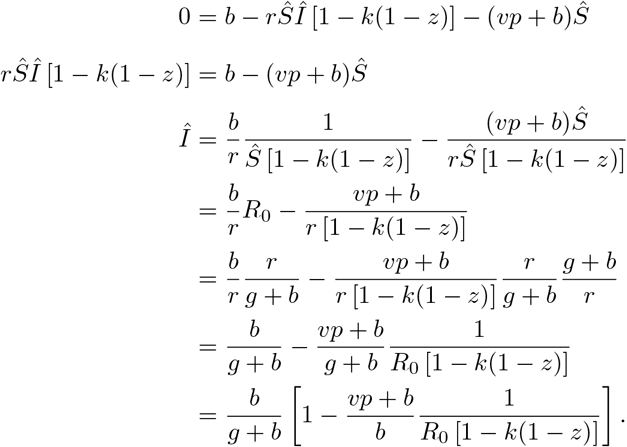

We still need to know 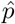 and 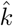, which require knowing the sentiment equilibria.

### A.2 Sentiment Subsystem Equilibria

If we track the sentiment marginal variables *PN, PM*, and *AN* (noting that *AM* = 1 − *PN* − *PM* − *AN*), we obtain

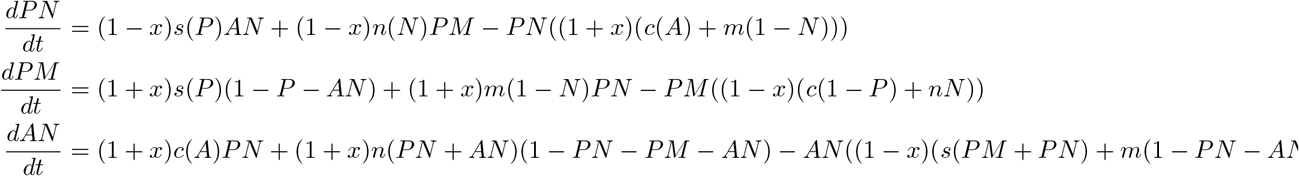

From this system of equations, it is not obvious what the equilibrium values of these variables are. Verifying that all the boundaries where either one or none of the three variables is 1, and the rest are zero, are indeed equilibria, requires only inspection. We label these equilibra by 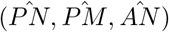.

The Jacobian of this system is

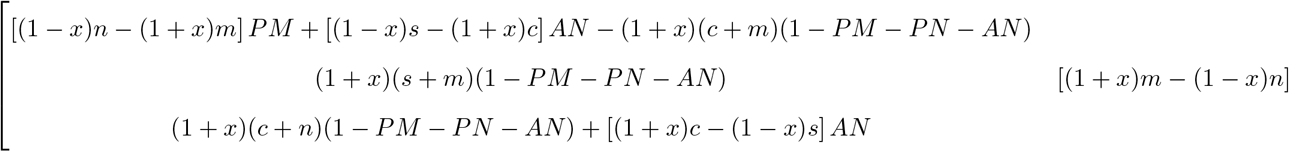

The eigenvalues of the equilibrium (1, 0, 0) (which corresponds to 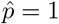 and 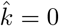 are:

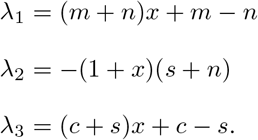

This equilibrium is stable when:

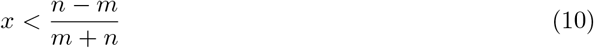

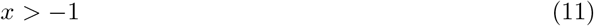

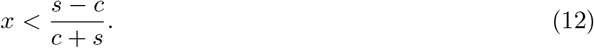

The eigenvalues of the equilibrium (0, 1, 0) (which corresponds to 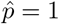 and 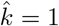 are:

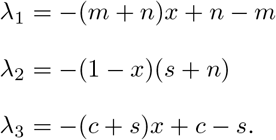

This equilibrium is stable when:

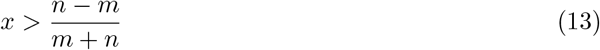

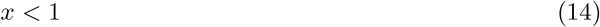

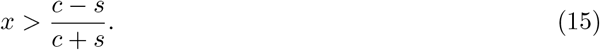

The eigenvalues of the equilibrium (0, 0, 1) (which corresponds to 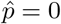 and 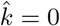 are:

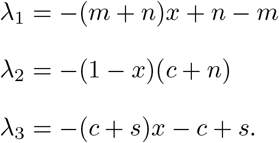

This equilibrium is stable when:

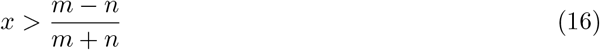

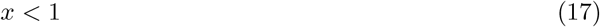

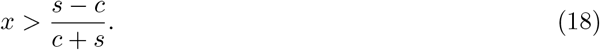

Finally, the eigenvalues of the equilibrium (0, 0, 0) (which corresponds to 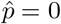 and 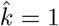 are:

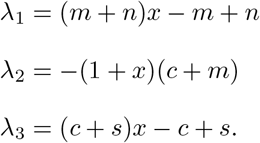

This equilibrium is stable when:

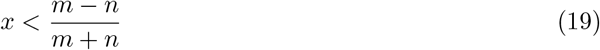

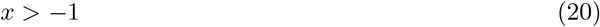

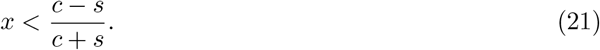

Internal equilibria only exist when specific transmission parameter values are exactly equal and so are irrelevant for real-life situations. We will not discuss them here.

### A.3 Disease Subsystem Stability

Because we are only concerned with situations, where 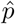 and 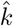 are either 0 or 1, we can now finish our analysis of the disease subsystem.

The Jacobian of the disease subsystem is

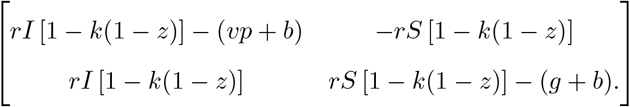

When *Î* = 0, the eigenvalues are

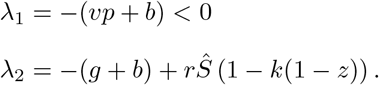

When 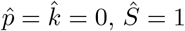 and

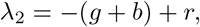

so this equilibrium is stable when *R*_0_ *<* 1.

When 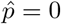 and 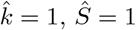and

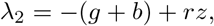

So this equilibrium is stable when 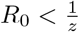.

When 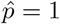 and 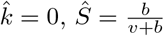 and

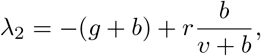

So this equilibrium is stable when 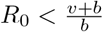

When 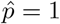 and 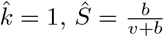 and

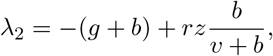

So this equilibrium is stable when 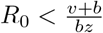

As is typical of these simple coupled SIR models, the reverse condition (i.e. when *R*_0_ is greater than one of these critical values), corresponds to the equilibrium with matching sentiment state but with *Î >* 0. See (14) for an example. Because this analysis is tedious and does not add anything mathematically to our understanding of the system, it is omitted here.

### A.4 Effects of Masking and Vaccination on Measles and Polio

When everyone is pro-vaccine (*p* = 1) and pro-mask (*k* = 1), the critical condition for endemic disease is

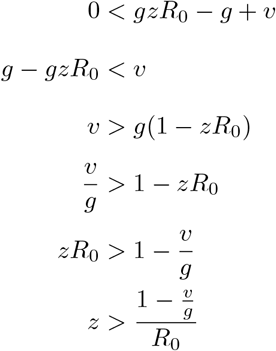

### A.5 Condition for No Epidemic Peaks

The condition for no epidemic peaks means that *I* has to monotonically decrease, so 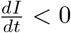 for all *t*. Provided that *I >* 0, we have

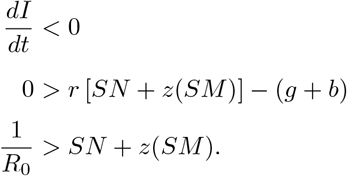

Our initial condition leads to

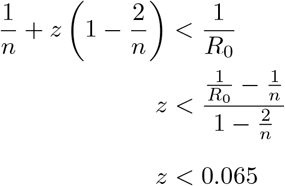

For an arbitrary initial susceptible fraction *S*_0_, we have

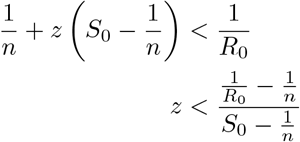

